# Changes of LipoxinA_4_ Levels Following Early Hospital Management of Patients with Non-Severe COVID-19: A Pilot Study

**DOI:** 10.1101/2022.04.18.22273880

**Authors:** Farzaneh Jamali, Bita Shahrami, Amirmahdi Mojtahedzadeh, Farhad Najmeddin, Amir Ahmad Arabzadeh, Azar Hadadi, Mohammad Sharifzadeh, Mojtaba Mojtahedzadeh

## Abstract

LipoxinA_4_ (LXA_4_) is an anti-inflammatory biomarker participating in the active process of inflammation resolution, which is suggested to be effective on infectious and inflammatory diseases like COVID-19. In this study, we hypothesized that LXA_4_ levels may increase following COVID-19 treatment and are even more accurate than commonly used inflammatory markers such as erythrocyte sedimentation rate (ESR), c-reactive protein (CRP), and ferritin. To test this hypothesis, a pilot study was conducted with 31 adult hospitalized patients with non-severe COVID-19. LXA_4_ levels were measured at the baseline and 48-72 hours later. Accordingly, ESR and CRP levels were collected on the first day of hospitalization. Moreover, the maximum serum ferritin levels were collected during the five days. LXA_4_ levels significantly increased at 48-72 hours compared to the baseline. ESR, CRP, and ferritin levels were positively correlated with the increased LXA4. In contrast, aging was shown to negatively correlate with the increased LXA_4_ levels. LXA_4_ may be known as a valuable marker to assess the treatment response among non-elderly patients with non-severe COVID-19. Furthermore, LXA_4_ could be considered as a potential treatment option under inflammatory conditions. Further studies are necessary to clarify LXA_4_ role in COVID-19 pathogenesis, as well as the balance between such pro-resolving mediators and inflammatory parameters.

## Introduction

The current coronavirus belongs to the Coronaviridae family which causes disease in human beings. It firstly emerged in Wuhan, China, but rapidly spread worldwide afterward and extended to a life-threatening pandemic. The virus was named SARS-CoV-2, which causes the coronavirus disease 2019 (COVID-19) [1]. The virus has some spike proteins on its outer surface that are necessary for binding to host cell receptors. When the virus enters the body, the immune system is activated, and the inflammatory processes then begin. Clinical manifestations of viremia can be ranged from asymptomatic to mild, moderate, severe, and critical diseases with exaggerated inflammation and cytokine storm. In severe cases of COVID-19, an uncontrolled inflammatory response with elevated cytokines and chemokines levels can result in multiple organ failures and worse patient’s outcomes [2].

Inflammation is an active process associated with the anti-inflammatory mediators’ production [3]. These mediators, called specialized pro-resolving mediators (SPMs), are a group of bioactive lipids (BALs) that resulted from arachidonic acid and other polyunsaturated fatty acids metabolisms. Lipoxins (LXs), maresins, protectins, and resolvins are SPMs involved in the active phase of the resolution, which is thought to control inflammation, promote healing, and limit tissue damage. LXs help in the recruitment of neutrophils to the site of inflammation and then in promoting neutrophils’ apoptosis to inhibit their excessive accumulation. Accordingly, this biomarker plays a role in the resolution process by delaying the apoptosis of macrophages as well as reducing the secretion of interleukin-8 [4]. It is believed that a complex correlation exists between inflammatory and non-inflammatory mediators in several inflammatory and infectious diseases, including COVID-19 [5-7]. The data on the role of such mediators in COVID-19 and their exact roles in the treatment and prevention of the disease is still lacking. Up to now, no specific biomarkers have been identified to assess the clinical response in patients with a mild to moderate disease. Moreover, the common biomarkers used to detect the severity of COVID-19 such as erythrocyte sedimentation rate (ESR), c-reactive protein (CRP), and ferritin, are not of high prognostic values [8, 9].

The principal aim of this study was to declare the trend of LXA_4_ changes in non-severe COVID-19 as well as its correlation with some commonly used inflammatory markers.

## Materials and Methods

### Study design and participants

This observational pilot study was performed at a tertiary university-affiliated hospital in Iran. The study protocol was approved by the ethics committee of Sina Hospital and Tehran University of Medical Sciences (TUMS) (IR.TUMS.MEDICINE.REC.1399.755). Patients’ informed consent was obtained before enrolment. The adult patients hospitalized due to non-severe COVID-19 were enrolled in this study. COVID-19 was ascertained by the presence of at least one of the related clinical symptoms such as fever, cough, and myalgia, besides a positive reverse transcriptase-polymerase chain reaction (RT-PCR) test or pulmonary computerized tomography (CT) scan findings indicating the hospital admission. On the other hand, the non-severe disease was defined as the absence of severe hypoxia (oxygen saturation less than 90%) and lung involvement (more than 50%). The patients received the hospital treatment protocol for COVID-19, including oxygen supplements, oral antiviral drugs, and symptoms’ treatment. Of note, those patients who received investigational agents on clinical trials were excluded from the study. The demographic and physiological parameters in this study were collected using the Hospital Information System (HIS) and patients’ records. These patients were followed-up for five days.

### LXA_4_ measurement

Serum samples for measuring LXA_4_ levels were collected once at the time of admission (LX0) and once 48-72 hours later (LX1) for each patient. At first, three milliliters of blood samples were allowed to clot for 10-15 minutes at room temperature. The obtained samples were then centrifuged (at 2000-3000 RPM) for 10 minutes. Thereafter, the supernatants were carefully collected and then stored at -70° C until collecting all the samples. LXA_4_ levels were measured using the biotin double antibody sandwich technology via a human ELISA kit in terms of the manufacturer’s instructions. The human LXA_4_ ELISA kit was purchased from German company Zellbio (Cat. No: ZB-10612C-H9648, Lot. No: ZB-OEH50820610-063). LXA_4_ levels are represented as nanogram per liter (ng/L) of serum. The assay range was from 75 to 2400 ng/L, and the sensitivity was measured as 9.5 ng/L. Therefore, LXA_4_ levels below the detection value (<9.5 ng/L) were considered as 9.5 ng/L for the statistical analysis.

### Inflammatory markers measurement

ESR and CRP levels and the maximum serum ferritin were collected on the first day of hospital admission and during the hospitalization, respectively.

### Statistical Analysis

Statistical analyses were performed using Kolmogorov–Smirnov, Wilcoxon Signed-Ranks, Mann-Whitney, and Spearman coefficient tests. As well, Statistical Package for Social Sciences (SPSS), version 24 was used. The level of statistical significance was defined as *P*-value <0.05.

## Results

Fifty-six patients were enrolled in the present study, but 25 patients were excluded based on the following exclusion criteria: early death (4 patients), early discharge (16 patients), and receiving investigational drugs (5 patients). Finally, the data obtained from a total of 31 patients were analyzed. A summary of the patients’ characteristics and demographics is represented in Table 1. The mean (±SD) age of the patients, including 18 men and 13 women, was 61.9±17 years old.

**Table 1.** Baseline demographic and clinical characteristics of patients

| Parameter | Number (%) |
| --- | --- |
| **Age (years old)** |  |
| 18-59 | 11 (35.5%) |
| 60-74 | 12 (38.7%) |
| 75-85 | 8 (25.8%) |
| **Gender** |  |
| Male/Female | 18/13 (58%/42%) |
| **Body weight (kilogram) (Mean±S.D)** | 77±14.3 |
| **Signs and symptoms on admission** |  |
| Fever | 8 (25.8%) |
| Cough | 18 (58%) |
| Chills | 12 (38.7%) |
| Myalgia | 15 (48.3%) |
| Gastrointestinal problems | 15 (48.3%) |
| Dyspnea | 19 (61.2%) |
| **PCR test result** |  |
| Positive | 13 (42%) |
| **Pharmacologic treatment following hospital stay** |  |
| Hydroxychloroquine | 25 (80%) |
| Lopinavir/Ritonavir | 21 (68%) |
| Umifenovir | 14 (45%) |
| Non-steroidal anti-inflammatory drug (NSAID) | 0 (0%) |
| Corticosteroid | 6 (19%) |

Changes of LXA_4_ levels are shown in Fig. 1. In 29% of the studied patients, LXA_4_ levels remained below 9.5 ng/L, of which 78% were elderly, LXA_4_ decreased in 3%, and there was an increase in LXA_4_ levels in 68% of the patients compared to baseline concentrations. LXA_4_ levels at the time of admission and 48-72 hours later were obtained as 9.5 [9.5, 9.9] and 16.1 [9.5, 25.4] ng/L, respectively. LXA_4_ concentrations significantly increased after early interventions in the hospital (*P*<0.05). In addition, there was an inverse correlation between age and changes in LXA_4_, indicating that LXA_4_ levels less increased with aging (*R*= -0.375; *P*=0.037). A subgroup analysis by aging is also shown in Fig. 1. The mean baseline LXA_4_ was calculated as 9.87±0.74 ng/L in the elderly (≥65 years old) and 10.03±0.82 ng/L among the non-elderly patients (<65 years). LXA_4_ concentrations increased to 16.23±10.10 ng/L in the elderly group and more than doubled in the non-elderly group during 48-72 hours. No significant difference was found in LXA_4_ changes in terms of the patients’ gender (*P*=0.825).

**Fig. 1:**
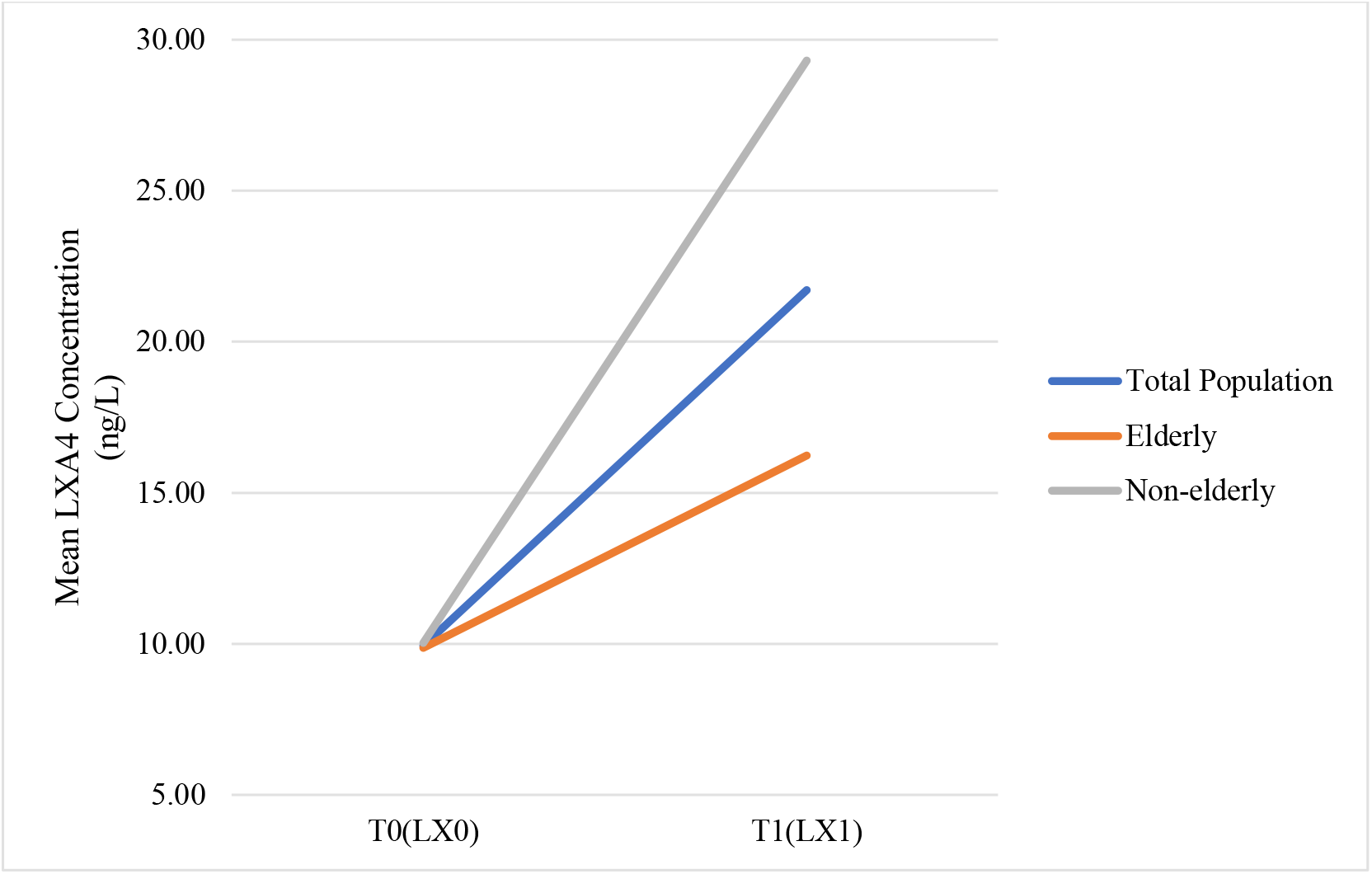
LXA_4_ level changes by age. The LXA_4_ levels increased in 48-72 hours, and the highest increase occurred in the non-elderly population.

The mean (±SD) baseline concentrations of ESR, and CRP as well as the maximum concentration of ferritin were 55.7±33.7 mm/h, 74.7±57.3 mg/L, and 568.7±530.0 ng/mL, respectively. The results show that the patients with higher serum ESR and CRP levels at the time of admission also had a greater increase in LXA_4_ concentration (*R*=0.535, 0.499; *P*=0.005, 0.007; respectively) (Fig. 2 and 3). The positive correlation between the maximum ferritin levels and the LXA_4_ changes was statistically significant as well (*R*=0.398; *P*=0.043) (Fig. 4).

**Fig. 2:**
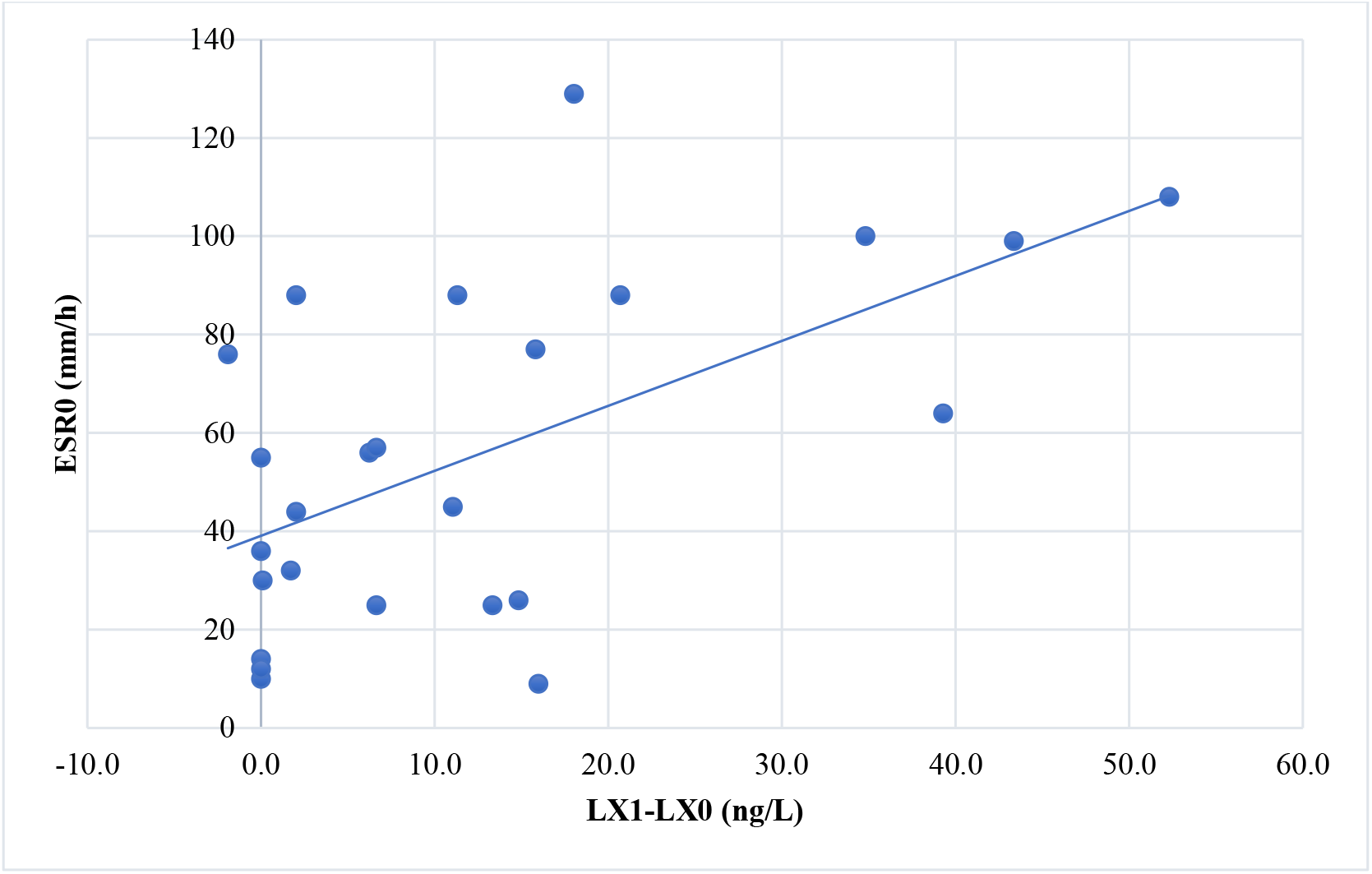
The correlation between ESR_0_ and changes of LXA_4_

**Fig. 3:**
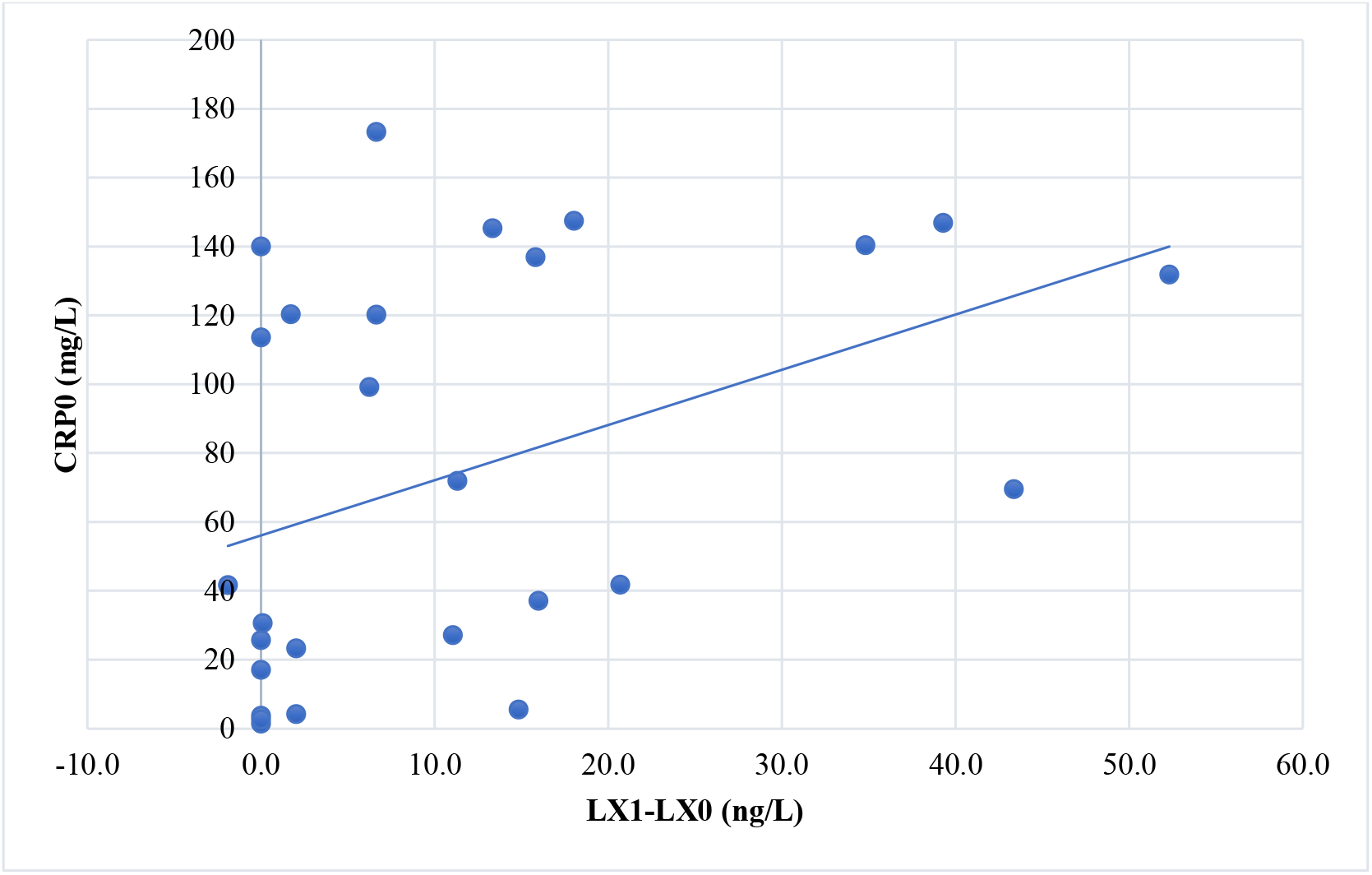
The correlation between CRP_0_ and changes of LXA_4_

**Fig. 4:**
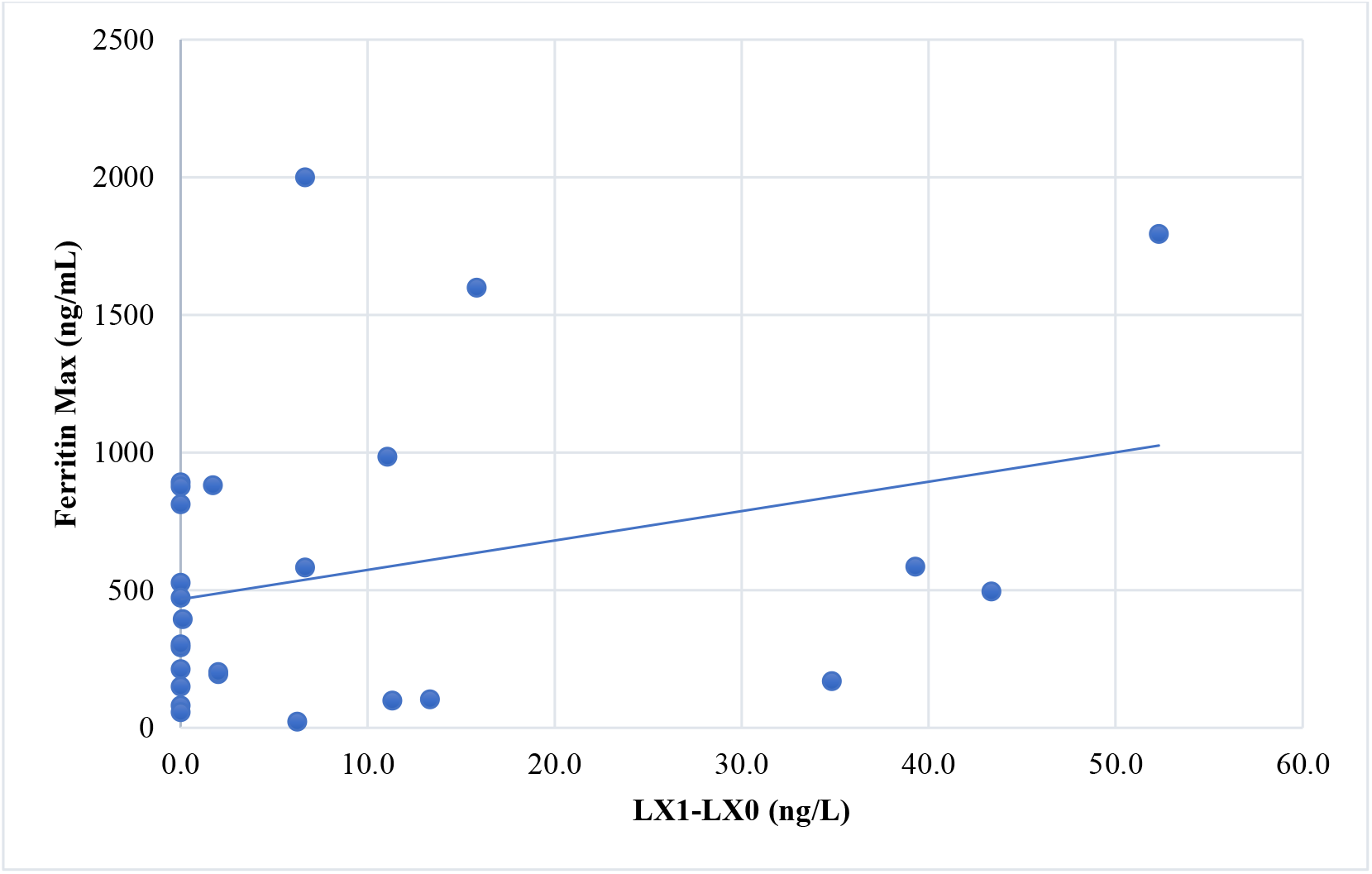
The correlation between maximum ferritin and changes of LXA_4_

## Discussion

Inflammation is an essential host immune response to pathogens, followed by some active resolution processes to restore homeostasis. LXs are anti-inflammatory mediators that increase at early stages of resolution and promote tissue regeneration, neutrophils apoptosis, macrophage phagocytosis, and cytokine reduction along with other specialized pro-resolving mediators [10]. Despite conducting extensive research on the pro-inflammatory mediators, the pro-resolving roles in COVID-19 have been poorly studied. To the best of our knowledge, this is the first clinical study evaluating serum LX changes among patients with COVID-19. This pilot study showed that LXA_4_ levels increased in the hospitalized patients with non-severe COVID-19 following performing the early therapeutic interventions.

Yang et al. in their study [11] have reviewed the role of SPMs in pulmonary diseases associated with inflammation. They have concluded that LXs and other SPMs have protective effects on the bacterial and viral invasions of the lungs by inhibiting the replication of microorganisms as well as regulating the immune system. Additional studies [12, 13] have shown a significant correlation between severe asthma and the reduced levels of LXA_4_ in serum and bronchoalveolar lavage fluid (BALF). Furthermore, LXs levels significantly decreased in sputum samples of chronic obstructive pulmonary disease (COPD) patients [14]. The imbalance between inflammatory factors results in hypo-differentiation to M2 macrophages, the accumulation of apoptotic cells, and progressive fibrosis in the airways [11].

The role of SPMs in infectious diseases was reviewed in a study by Shayna Sandhaus and Andrew G. Swick [15]. In animal models of sepsis, LXA_4_, Maresin R_1_, and resolvins by suppressing the NF-κB pathway effectively reduced systemic inflammation, bacterial burden, and pro-inflammatory mediators’ production [16-18]. SPMs also showed some positive effects on treating sepsis, improving survival, and reducing the need for antibiotics [19]. In the context of the influenza virus, SPMs in preclinical studies were found to be inversely correlated with both the severity and dissemination of more virulent strains [20]. In addition, SPMs like LXB_4_ showed potential properties in enhancing the immunity of the influenza vaccine by increasing antibody titers through the B cell activation [21]. While previous studies have shown great promise in improving outcomes in the field of infectious and inflammatory diseases, some suggest that the resolution’s timing is a paramount factor playing a role in establishing a proper inflammatory response [15]. In a mouse model of pneumosepsis, LXA_4_ treatment at the early stages of sepsis worsened the outcomes, while at the later stages, it improved survival and inflammation [22]. However, other studies showed the beneficial effects of early administration of SPMs on sepsis [15]. In tuberculosis infection, a sharp increase was observed in pro-inflammatory and anti-inflammatory mediators in the acute phase of disease in mice, where exogenous SPM could impair the balance between inflammatory properties and the Th1-mediated immune response [23]. However, the timely administration of SPMs could reduce TNF-α production and bacterial growth in human macrophages [24].

There are reports of the involvement of lipid mediators in COVID-19, stating that when human cells are exposed to SARS-CoV-2 and the human coronavirus 229E (HCoV-229E), they secrete large amounts of BALs, which then inactivate the virus [25, 26]. Yan B et al. in their study [25] have found that administering linoleic acid and arachidonic acid to Huh-7 cells infected with HCoV-229E and the Middle East Respiratory Syndrome Coronavirus (MERS-CoV) could interfere with virus replication. Additionally, Anne-Sophie Archambault et al. [27] have assessed the lipid storm in severe cases of COVID-19 by measuring the BALs concentrations in BALF. Most SPMs are detectable in the BALF of patients, among which LXA_4_ is the most significant one. In addition, the levels of SPMs and pro-inflammatory lipids such as prostaglandins (PG) and leukotrienes, were simultaneously high, which indicated the coexistence of such mediators in the acute phase of inflammation, while the resolution process was not fully engaged. LXA_4_ was indicated to have a significant correlation with PGE_2_, PGD_2_, and thromboxane B_4_, but it had no significant correlation with clinical parameters and aging. In the present study, in contrast, the changes of LXA_4_ were observed to be correlated with some of the clinical parameters of patients. The increased ESR, CRP, and ferritin were demonstrated to be associated with higher inflammation and more severe disease [28]. The positive correlation of LXA_4_ with the baseline ESR and CRP levels as well as the maximum ferritin levels in the present study indicated that the patients with higher inflammatory states secreted more LXA_4_. Correspondingly, this suggests that in a healthy inflammatory process, pro-inflammatory and pro-resolving phases are two sides of the same coin. These results are consistent with previous data presenting that LXA_4_ elevation occurs due to polyunsaturated fatty acid metabolism in the acute phase of inflammation [27, 29]. As well, it has been suggested that the release of arachidonic acid occurs in two phases in inflammatory events. Firstly, arachidonic acid is metabolized to PGE_2_ as an inflammatory mediator, anti-inflammatory processes are initiated, and then LXA_4_ production increases [30]. However, the exact time of this conversion is not fully known yet, and more studies are needed to determine the precise position of LXs in inflammatory processes. Corticosteroids have been shown to suppress the production of LXs along with pro-inflammatory mediators [31, 32]. Corticosteroids appear to be effective in severe cases of COVID-19 in which both excessive inflammatory response and cytokine storm occur [31]. However, early and high doses of corticosteroids in non-severe cases could not be helpful [31], which may be due to their effect on pro-resolving roles.

In this study, another finding was the significant inverse correlation between aging and changes in LXA_4_ concentration. The effects of aging factors such as immunosenescence and inflammaging on the COVID-19 outcomes have been previously investigated in many studies [33-35]. The baseline sterile inflammation caused by the increased cytokines produced by senescent cells [36] predisposes the elderly to hyper-inflammatory response and impaired resolution. This hypothesis is consistent with our results, revealing that among elderly patients, the increase in LXA_4_ was less during early days of the treatment.

The present study results suggest that along with the inflammatory response to pathogens, active resolution, and the timely conversion to resolution phase are of great importance in having an effective fight on the coronavirus. Therefore, LXA_4_ and other SPMs without any immunosuppressive effect may be potential treatment options under inflammatory conditions. Besides, early changes of LXA_4_ in our study showed that such mediators might be valuable markers for assessing the treatment response compared to common-used inflammatory markers with fewer changes. However, the LXA_4_ use is not recommended in the elderly population, as the baseline LXA_4_ levels were below the detection cut-off in many of them and did not significantly increase compared to the non-elderly patients.

Although less information is currently available on the possible role of LXA_4_ following infecting with COVID-19, the present study investigated the trend and pattern of early changes of serum LXA_4_ among the hospitalized COVID-19 patients. However, this pilot study also had some limitations. Firstly, the small sample size made it difficult to interpret our findings. The second limitation is the lack of a control group. Next, we did not explore the factors affecting LXA_4_ serum levels, including the onset of symptoms in patients, comorbidities, and medication history. Moreover, we did not exclude the patients receiving anti-inflammatory drugs (e.g., corticosteroids) before the second sampling.

## Conclusion

LXA_4_ is suggested to be a beneficial biomarker in infectious and inflammatory diseases like COVID-19. This pilot study showed that LXA_4_ increased shortly after therapeutic interventions among hospitalized patients with non-severe COVID-19. Moreover, it was significantly correlated with patients’ aging and inflammatory markers. Further studies are needed to clarify the role of LXA_4_ in the pathogenesis of COVID-19 as well as the balance between inflammatory and anti-inflammatory mediators.

## Data Availability

All data produced in the present work are contained in the manuscript

## Data Availability

All data generated or analyzed during this study are included in this published article.

## Acknowledgements

The research was previously presented as an abstract at the 6^th^ international conference on prevention & infection control (ICPIC 2021).

